# Impact of Geographic and Person-Centered Barriers on Clinical Outcomes of Latino Patients With Multiple Sclerosis and Related Disorders

**DOI:** 10.64898/2026.05.29.26354488

**Authors:** Lily Finkelstein, Paola Rosario, Alejandro Martinez, Irena Dujmovic Basuroski, Deanna Saylor, Monica M. Diaz

## Abstract

**Background:** Social and geographic barriers contribute to worse outcomes in patients with multiple sclerosis (MS) and related disorders, but these factors remain poorly characterized among Latino patients. We evaluated associations between distance to specialty care, neighborhood deprivation, insurance status, and clinical outcomes among Latinos with MS and related disorders.

**Methods:** We conducted a retrospective study of Latino adults with MS, neuromyelitis optica spectrum disorder, and myelin oligodendrocyte glycoprotein antibody-associated disease. Demographic, clinical, and socioeconomic variables were abstracted from the medical record. Distance to care was defined as residence ≥50 vs. <50 miles from clinic and neighborhood deprivation as Area Deprivation Index (ADI) state rank. We used unadjusted and multivariable regression to evaluate associations with Expanded Disability Status Scale (EDSS) score, annualized relapse rate (ARR), and disease-modifying therapy (DMT) non-adherence.

**Results:** Among 99 Latino patients, 84 had MS, 11 MOGAD, and 4 NMOSD; 46.5% lived ≥50 miles from clinic. Living ≥50 miles from clinic was associated with higher EDSS scores in unadjusted analyses, but not after covariate adjustment. In multivariable analyses, Medicaid insurance was associated with higher EDSS compared with commercial insurance (β=1.071, p=0.031) and higher ARR (β=0.230, p=0.022). Higher ADI showed a non-significant trend toward higher EDSS (β=0.147 per 1-decile increase, p=0.068). DMT non-adherence was not significantly associated with covariates.

**Conclusions:** In this cohort of Latinos with CNS demyelinating diseases, Medicaid insurance was associated with greater disability level and higher relapse activity. These findings suggest that insurance status should be considered when designing strategies to improve access to neuroimmunology care.

## INTRODUCTION

Multiple sclerosis (MS) is the most common chronic inflammatory demyelinating disease of the central nervous system (CNS) affecting an estimated 900,000 individuals in the United States(Qian et al., 2023; Walton et al., 2020). Although less common, neuromyelitis optica spectrum disorder (NMOSD) and myelin oligodendrocyte glycoprotein associated disorder (MOGAD) are related immune-mediated demyelinating disorders that can lead to significant neurologic disability(Carnero Contentti and Correale, 2021; Sechi et al., 2022). Epidemiologic studies suggest that the burden and clinical course of CNS demyelinating disease may vary by race and ethnicity(Dykes et al., 2024; Mercado et al., 2020; Petracca et al., 2023), though Latino/a/x (herein referred to as Latino) individuals remain underrepresented in MS and neuroimmunology studies contributing to a significant gap in knowledge on clinical outcomes of Latinos with CNS demyelinating diseases(Langer-Gould et al., 2022; Moore et al., 2023). This is crucial to investigate, as it is estimated that the number of Latino people with MS is expected to rise from 14% in 2005 to 29% by 2050(Amezcua et al., 2017).

Adverse social and geographic factors may negatively impact clinical outcomes in CNS demyelinating diseases through heterogeneous factors, including delayed diagnosis, reduced access to subspecialty care, interruptions in disease-modifying therapy (DMT), and barriers to longitudinal monitoring. These factors are particularly relevant in MS, NMOSD, and MOGAD, where timely diagnosis, sustained treatment adherence, relapse recognition, MRI surveillance, and ongoing neurology follow-up are needed to reduce disease activity and disability accumulation(Amezcua et al., 2021; Dobson et al., 2022; Hjerthen et al., 2024). Prior studies have found associations between lower socioeconomic status, lower health literacy, insurance-related barriers, and neighborhood-level socioeconomic disadvantage with worse outcomes among people with MS, including greater disability accrual, reduced access to DMTs, lower DMT adherence, and higher healthcare utilization(Abbatemarco et al., 2022; Boorgu et al., 2022; He et al., 2023; Marrie et al., 2014; Simacek et al., 2018). However, fewer studies have examined these relationships specifically within Latino populations with MS and related demyelinating disorders.

Neighborhood-level deprivation may be an underrecognized factor affecting healthcare access-related differences in CNS demyelinating disease outcomes(Abbatemarco et al., 2022; Boorgu et al., 2022). The Area Deprivation Index (ADI) is a validated area-level measure of socioeconomic disadvantage that incorporates indicators related to income, education, employment, and housing quality(Kind and Buckingham, 2018). Distance to specialty care and insurance status may also contribute to clinical outcomes in patients with CNS demyelinating disease. Patients living farther from tertiary neuroimmunology clinics may face transportation barriers, travel costs, time away from work, and difficulty attending urgent visits for new symptom or relapse evaluations (Lin et al., 2023; Solomon et al., 2021; Turner et al., 2013). Insurance status may also impact clinical outcomes. Prior studies have shown that insurance coverage affects DMT use, and that insurance restrictions can delay treatment initiation and contribute to increased disease activity in MS (Mizell, 2024; Wang et al., 2016). Understanding how these factors relates to outcomes within Latino patients may help identify modifiable barriers and improve care for Latino people with CNS demyelinating diseases. Therefore, we evaluated Latino adults with MS, NMOSD, or MOGAD receiving care at the University of North Carolina MS/Neuroimmunology clinics and examined the associations between distance to specialty care, neighborhood deprivation measured by ADI, insurance status and clinical outcomes.

## METHODS

### Study Design and Setting

We conducted a retrospective cohort study of Latino adults with an ICD-10 code diagnosis of MS(Montalban et al., 2025; Thompson et al., 2018), NMOSD(Wingerchuk et al., 2006), or MOGAD(Banwell et al., 2023) based on the electronic health record who received care at the University of North Carolina (UNC) MS/Neuroimmunology clinics between February 1, 2020 and February 28, 2026. Data were extracted from the patient’s electronic medical record and entered into REDCap by two research assistants (LF, PR) with adjudication and review of imaging variables by a study neuroimmunoloigst (MMD).

### Participants

Eligible participants were adults aged ≥18 years who self-identified as Latino or Hispanic in the electronic medical record, had an established diagnosis of MS, NMOSD, or MOGAD assigned in their electronic medical record chart by their treating neurologist, and had at least one clinical encounter in the UNC MS/Neuroimmunology clinics during the study period. Patients were excluded if chart documentation was insufficient to confirm diagnosis or determine key outcomes, including Expanded Disability Status Scale (EDSS), relapse history, or DMT adherence.

### Patient Identification and Data Abstraction

Potentially eligible patients were identified using TriNetX queries based on meeting all of the following criteria: (1) Hispanic/Latino ethnicity; (2) diagnosis codes for MS, NMOSD, or MOGAD; (3) having at least one encounter at a UNC MS/Neuroimmunology clinic location, and (4) encounter dates within the specified study period. In the case there were multiple encounter dates within the study period, the most recent encounter from the date of data collection was used for data abstraction, or the encounter that was deemed to have the most complete set of key exposure and outcome variables. After the cohort was identified through TriNetX, a list of medical record numbers was generated by the Carolina Data Warehouse for Health that allowed us to identify patients’ medical records. Two research assistants abstracted variables from the neuroimmunology office visit note including age at the time of the office visit, sex, diagnosis, disease phenotype (relapsing, monophasic, progressive), date of diagnosis, disease duration, type, residential 9-digit ZIP code, vitamin D supplementation, DMT use, self-reported DMT adherence, number of relapses reported over the disease course, MRI brain and spinal cord outcomes (new or enlarging T2 lesions, gadolinium-enhancing lesions noted within the study period), and the EDSS recorded at the time of the most recent visit from the time of data collection. For all MRI and EDSS chart variables, one study neuroimmunologist (MMD) reviewed the data extraction outcomes to ensure quality of the data and accuracy. Data abstraction followed a standardized chart review protocol.

### Exposures and Covariates

Key demographics were extracted including age, sex, insurance type (Medicare, Medicaid, commercial insurance, or none/Charity Care), and residential zip code. In the case of dual insurance, if Medicare was the primary insurance, we considered Medicare for insurance status. Zip code was used to calculate the patient’s distance from the UNC MS Clinic using Google Maps and to approximate neighborhood-level socioeconomic factors, dichotomized as ≥50 versus <50 miles, chosen based on prior literature using similar cut-offs for travel distance from neurology clinics (Lin et al., 2023; McGinley et al., 2024). Residential ZIP code was also linked to the ADI, a validated neighborhood-level measure of socioeconomic disadvantage using the University of Wisconsin ADI database(Kind and Buckingham, 2018). ADI national rank is a percentile measure that ranks neighborhoods relative to all neighborhoods across the United States. ADI national rank ranges from 1 to 100, with higher values indicating greater neighborhood deprivation based on factors related to income, education, employment, and housing quality. ADI state rank ranges from 1 to 10, with higher values indicating greater neighborhood deprivation relative to other areas within the same state. Covariates included age, sex, disease duration, vitamin D supplementation at the time of the office visit, and disease phenotype, and relapses. Relapses were also dichotomized as no documented relapse vs. ≥1 documented relapse after disease onset.

### Outcomes

The primary clinical outcomes were most recent EDSS, annualized relapse rate (ARR), and self-reported DMT non-adherence noted in the neuroimmunology office visit note. EDSS was analyzed as a continuous variable and dichotomized as EDSS ≥5 vs. <5 to represent moderate to severe disability. EDSS is scored from 0 to 10 with greater numbers indicating greater disability. A cut-off of 5.0 indicates the patient can walk about 200 meters without aid or rest, but disability is severe enough to impair full daily activities(Kurtzke, 1983); scores above this threshold increasingly reflect ambulatory impairment, including need for unilateral or bilateral assistance and wheelchair dependence at higher levels. This threshold is clinically meaningful and has been used as in several MS clinical trials(Gold et al., 2012; Polman et al., 2006). ARR was calculated as the number of documented clinical relapses after disease onset divided by disease duration in years. DMT non-adherence was analyzed as a binary outcome and was defined using clinician documentation of at least one delayed or missed infusions/injection DMT or any documentation of non-adherence for oral DMTs, failure to initiate a prescribed DMT, or other chart evidence of inconsistent DMT use during the study period. DMT non-adherence analyses were restricted to participants who had a prescription for DMT at the office visit included in the study data abstraction.

### Statistical Analysis

Descriptive statistics were used to summarize demographic, clinical, and socioeconomic characteristics. Continuous variables were summarized using means and standard deviations or medians and interquartile ranges, as appropriate. Categorical variables were summarized using frequencies and percentages. EDSS and ARR were compared in subgroups by distance category using Mann-Whitney U tests because these outcomes were not normally distributed. Associations between categorical variables and DMT non-adherence were evaluated using Fisher’s exact test.

Multivariable linear regression models were used to evaluate associations between exposures and continuous outcomes, including EDSS and ARR. Logistic regression models were used to evaluate associations with DMT non-adherence. Because the number of DMT non-adherence events was small, the DMT non-adherence logistic regression model was considered exploratory. Models adjusted for clinically relevant covariates, including age, sex, disease duration, vitamin D supplementation, disease phenotype, distance category, ADI state rank, and insurance type. All analyses were conducted using Jamovi statistical software version 2.7 with p<0.05 considered significant.

### Ethical Considerations

This study was approved by the University of North Carolina Institutional Review Board (IRB#25-0510). The requirement for informed consent was waived because the study involved retrospective review of existing clinical data and posed minimal risk to participants. All data were abstracted and analyzed in accordance with institutional privacy and data security requirements. Protected health information was stored separately from the analytic dataset and accessed only by approved study personnel.

## RESULTS

During the study period, 2,250 adults with a diagnosis code of MS, MOGAD, or NMOSD were evaluated in the UNC MS/Neuroimmunology Division. Of these, 150 patients (6.7%) were noted in the electronic medical record to self-identify as Latino/Hispanic, and 99 met inclusion criteria for confirmed MS, MOGAD, or NMOSD.

Demographic, medical and neuroimmunologic characteristics are shown in **Table 1**. The mean age at visit was 39.9 years (SD 12.4), and 72 patients (72.7%) were female. Most patients had MS (84.8%), followed by MOGAD (11.1%) and NMOSD (4.0%). Relapsing-remitting disease was the most common phenotype (76.8%). The mean disease duration was 7.9 years (SD 6.6), the median number of prior DMTs was 1.0 (IQR 0.0–3.0), and the mean annualized relapse rate was 0.38 (SD 0.42). The mean distance to clinic was 70.1 miles (SD 96.6), and the mean ADI national rank was 60.8 (SD 22.5) and state rank was 5.6 (SD 2.7).

**Table 1.** Demographic and Clinical Characteristics of Patients with Demyelinating Diseases (N=99)

| **Characteristic** | **Mean (SD) or Median (IQR) or n (%)** |
| --- | --- |
| Age at visit | 39.9 (12.4) |
| Female sex | 72 (72.7%) |
| Self-reported Race |  |
| Other | 73 (73.7%) |
| White/Caucasian | 20 (20.2%) |
| Black/African American | 3 (3.0%) |
| American Indian/Native Alaskan | 3 (3.0%) |
| Latino/Hispanic Ethnicity | 99 (100%) |
| Insurance type |  |
| Commercial insurance | 39 (39.4%) |
| Medicaid | 26 (26.3%) |
| Medicare | 13 (13.1%) |
| None or Charity Care | 12 (12.1%) |
| Other insurance type (Tricare or other) | 9 (9.1%) |
| Neuroimmunologic Diagnosis |  |
| Multiple Sclerosis | 84 (84.8%) |
| MOGAD | 11 (11.1%) |
| NMOSD | 4 (4.0%) |
| Disease phenotype |  |
| Relapsing-remitting | 76 (76.8%) |
| Monophasic | 10 (10.1%) |
| Secondary progressive | 8 (8.1%) |
| Primary progressive | 5 (5.1%) |
| Disease duration (years) | 7.9 (6.6) |
| Number of prior DMTs | 1.0 (0.0, 3.0) |
| Total number of relapses during disease course | 1.0 (1.0, 2.5) |
| Annualized Relapse Rate | 0.38 (0.42) |
| EDSS | 2.0 (1.0, 4.5) |
| EDSS $\geq 5$ | 24 (24.2%) |
| Currently not on DMT | 14 (14.1%) |
| DMT non-adherence | 6/85 (7.1%) |
| On Vitamin D | 39 (39.4%) |
| Comorbidities or Non-Neurological Medical Characteristics |  |
| None | 32 (32.3%) |
| Hypertension | 19 (19.2%) |
| Smoking, current or former | 16 (16.2%) |
| Chronic lung disease | 13 (13.1%) |
| Current Pregnancy or pregnancy within the 1 year prior to office visit | 11 (11.1%) |
| Diabetes | 10 (10.1%) |
| Other neurologic condition | 3 (3.0%) |
| Social or Geographic Characteristics |  |
| Distance to clinic (miles) | 70.1 (96.6) |
| ≥50 miles | 46 (46.5%) |
| ADI national rank | 60.8 (22.5) |
| ADI state rank | 5.6 (2.7) |

**Abbreviations:** ADI = Area Deprivation Index; DMT = Disease-modifying therapy; IQR = Interquartile Range; MOGAD = Myelin oligodendrocyte glycoprotein antibody-associated disease; NMOSD = Neuromyelitis optica spectrum disorder; SD = Standard deviation

Clinical outcomes by distance to clinic are shown in **Table 2**. Patients living ≥50 miles from the clinic had higher EDSS scores than those living <50 miles away: median 3.0 (IQR 2.0–6.0) vs. 2.0 (IQR 1.0–2.5; p<0.001). EDSS distribution residential distance from the clinic is shown in **Figure 1**. ARR did not differ significantly by distance group. Among participants currently taking DMT, DMT non-adherence did not differ significantly by distance to clinic: 2/38 (5.3%) among those living ≥50 miles away versus 4/47 (8.5%) among those living <50 miles away (p=0.687).

**Figure 1.**
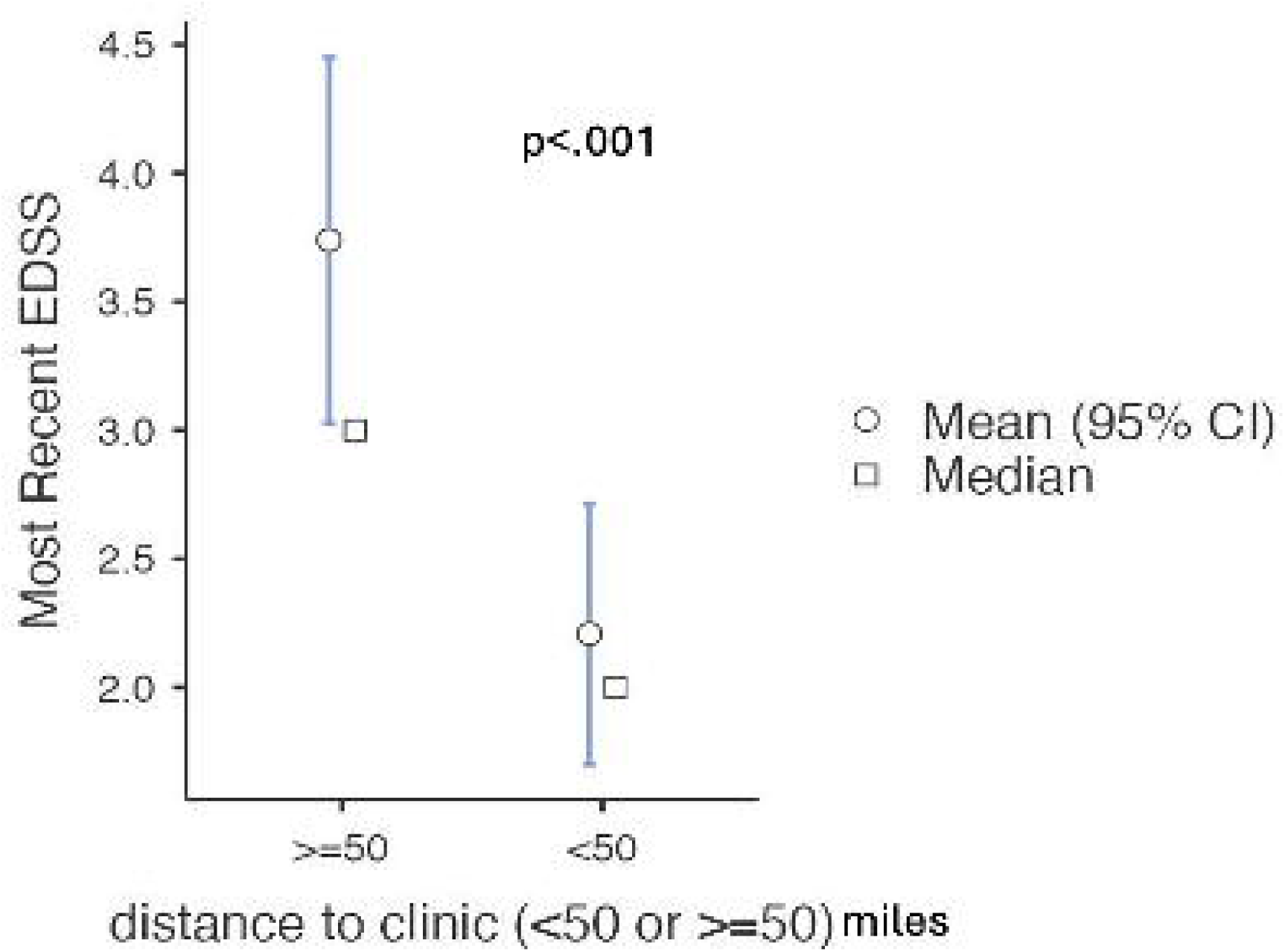
Most recent EDSS by distance to the UNC MS/Nauroimmunology clinic (N=99)

**Table 2.**
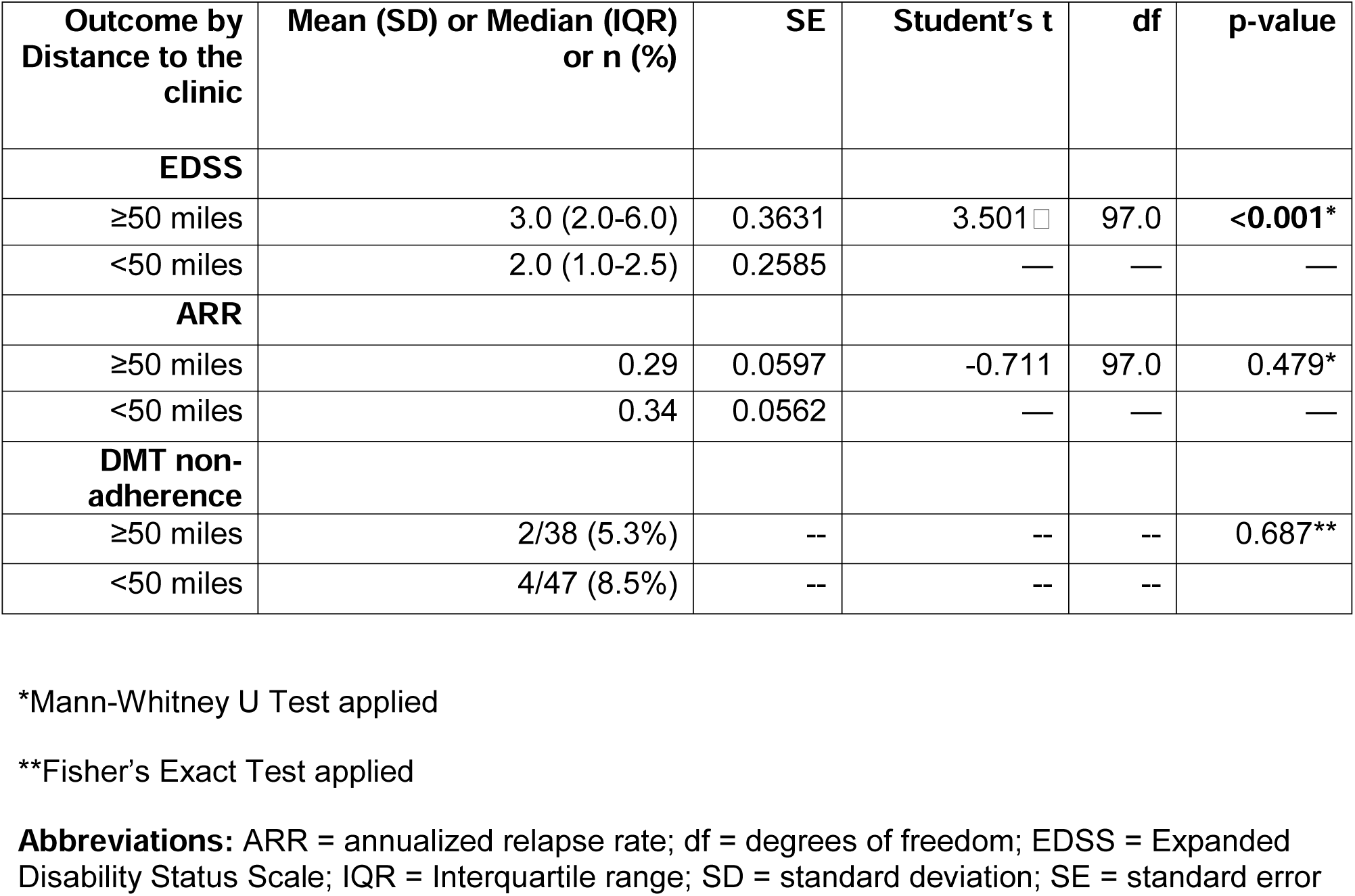
Clinical Outcomes of Latino adults with Demyelinating Diseases by distance to clinic (N=99)

| Outcome by Distance to the clinic | Mean (SD) or Median (IQR) or n (%) | SE | Student's t | df | p-value |
| --- | --- | --- | --- | --- | --- |
| **EDSS** |  |  |  |  |  |
| ≥50 miles | 3.0 (2.0-6.0) | 0.3631 | 3.501 | 97.0 | **<0.001\*** |
| <50 miles | 2.0 (1.0-2.5) | 0.2585 | — | — | — |
| **ARR** |  |  |  |  |  |
| ≥50 miles | 0.29 | 0.0597 | -0.711 | 97.0 | 0.479* |
| <50 miles | 0.34 | 0.0562 | — | — | — |
| **DMT non-adherence** |  |  |  |  |  |
| ≥50 miles | 2/38 (5.3%) | -- | -- | -- | 0.687** |
| <50 miles | 4/47 (8.5%) | -- | -- | -- |  |

\*Mann-Whitney U Test applied
\*\*Fisher's Exact Test applied
**Abbreviations:** ARR = annualized relapse rate; df = degrees of freedom; EDSS = Expanded Disability Status Scale; IQR = Interquartile range; SD = standard deviation; SE = standard error

In adjusted linear regression, older age, Medicaid insurance status, and disease phenotype were significant predictors of EDSS score (**Table 3**). Age was positively associated with EDSS (β=0.049, p=0.007), and those with Medicaid insurance had higher expected EDSS scores compared with commercial insurance (β=1.071, p=0.031). Relative to relapsing-remitting disease, secondary progressive disease (β=3.077, p<0.001) and primary progressive disease (β=2.041, p=0.017) showed higher expected EDSS scores, whereas monophasic disease showed lower expected scores(β=-1.477, p=0.018). ADI state rank showed a non-significant trend toward higher EDSS (β=0.147, p=0.068; **Figure 2**), while distance to clinic was not significantly associated with EDSS in multivariable analyses..

**Figure 2.**
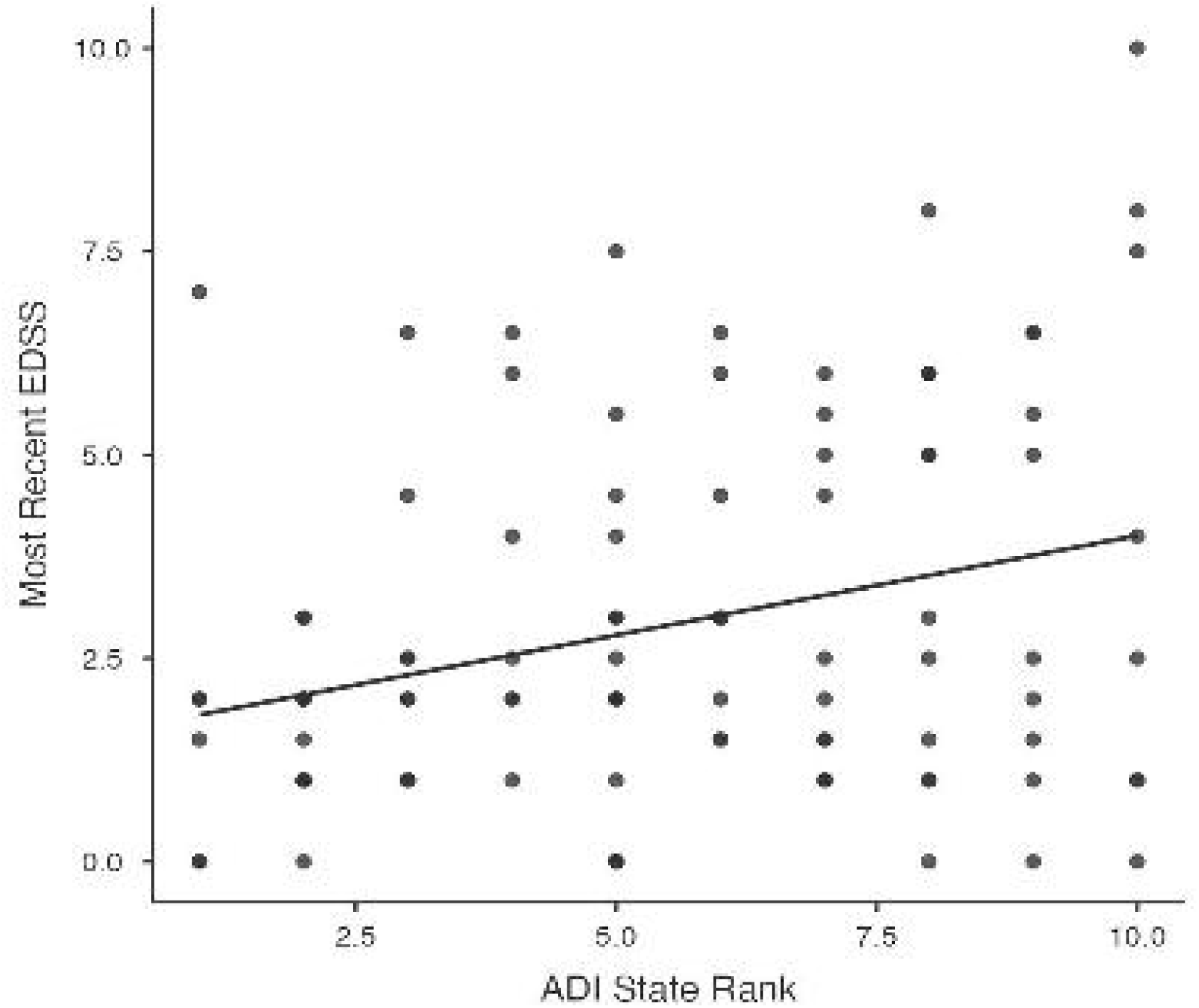
Most recent EDSS compared with ADI state rank among Latino adults with demyelinating disease(N=99)

**Table 3.** Multivariable linear regression analysis examining relationships between covariates and predictors of interest and Expanded Disability Status Scale (N=99)

| Predictor | Comparison / unit | $\beta$ estimate | p-value |
| --- | --- | --- | --- |
| Age | Per 1-year increase | 0.049 | 0.007* |
| Male sex | vs. female | 0.281 | 0.506 |
| No vitamin D supplementation | vs. yes | -0.053 | 0.888 |
| Disease duration | Per 1-year increase | 0.001 | 0.988 |
| ADI state rank | Per 1-decile increase | 0.147 | 0.068 |
| Distance to clinic <50 miles | vs. $\geq 50$ miles | -0.478 | 0.292 |
| Medicare insurance | vs. commercial insurance | 0.224 | 0.706 |
| Medicaid insurance | vs. commercial insurance | 1.071 | 0.031* |
| Other insurance | vs. commercial insurance | 0.061 | 0.929 |
| None/Charity Care | vs. commercial insurance | 0.018 | 0.975 |
| Monophasic disease | vs. relapsing-remitting disease | -1.477 | 0.018* |
| Secondary progressive disease | vs. relapsing-remitting disease | 3.077 | <0.001* |
| Primary progressive disease | vs. relapsing-remitting disease | 2.041 | 0.017* |

\*p-value<0.05
**Abbreviations:** ADI=Area Deprivation Index; SE=standard error. .

In adjusted linear regression for ARR as an outcome, longer disease duration was associated with lower ARR (β=-0.026, p<0.001), while Medicaid insurance status showed a higher expected ARR relative to commercial insurance status (β=0.230, p=0.022; **Table 4**). Patients not taking vitamin D at the time of the office visit had a higher ARR than those taking vitamin D (β=0.176, p=0.023). Compared with relapsing-remitting disease, monophasic disease (β=-0.492, p<0.001) and primary progressive disease (β=-0.340, p=0.048) showed a lower expected ARR. Distance to clinic and ADI state rank were not significantly associated with ARR.

**Table 4.** Multivariable linear regression examining relationships between covariates and predictors of interest and annualized relapse rate (N=99)

| Predictor | Comparison / unit | $\beta$ estimate | p-value |
| --- | --- | --- | --- |
| Age | Per 1-year increase | 0.003 | 0.452 |
| Male sex | vs. female | -0.015 | 0.861 |
| No vitamin D supplementation | vs. yes | 0.176 | 0.023 |
| Disease duration | Per 1-year increase | -0.026 | <0.001 |
| ADI state rank | Per 1-decile increase | 0.017 | 0.301 |
| Distance to clinic <50 miles | vs. $\geq 50$ miles | 0.116 | 0.203 |
| Medicare insurance | vs. commercial insurance | -0.014 | 0.905 |
| Medicaid insurance | vs. commercial insurance | 0.230 | 0.022 |
| Other insurance | vs. commercial insurance | 0.095 | 0.497 |
| None/Charity Care | vs. commercial insurance | -0.100 | 0.393 |
| Monophasic disease | vs. relapsing-remitting disease | -0.492 | <0.001 |
| Secondary progressive disease | vs. relapsing-remitting disease | -0.119 | 0.417 |
| Primary progressive disease | vs. relapsing-remitting disease | -0.340 | 0.048 |

**Abbreviations:** ADI=Area Deprivation Index

In multivariable logistic regression among patients who were prescribed a DMT at the time of the office visit, no covariates were significantly associated with documented DMT non-adherence (**Table 5**). Distance to clinic <50 miles was associated with higher but non-significant odds of DMT non-adherence compared with distance to clinic of ≥50 miles (OR 2.04, 95% CI 0.26–15.74; p=0.495). Disease duration was not significantly associated with DMT non-adherence (OR 1.07 per 1-year increase, 95% CI 0.92–1.24; p=0.399).

**Table 5.** Multivariable logistic regression analysis of non-adherence to disease-modifying therapy (n=85).

| Predictor | Comparison / unit | OR for DMT non-adherence | 95% CI | p-value |
| --- | --- | --- | --- | --- |
| Age | Per 1-year increase | 0.95 | 0.87-1.04 | 0.279 |
| Male sex | vs. female | 0.67 | 0.07-6.83 | 0.736 |
| Disease duration | Per 1-year increase | 1.07 | 0.92-1.24 | 0.399 |
| Distance to clinic <50 miles | vs. ≥50 miles | 2.04 | 0.26-15.74 | 0.495 |
| No vitamin D supplementation | vs. yes | 2.04 | 0.32-12.96 | 0.449 |

**Abbreviations:** CI = confidence interval; DMT = disease-modifying therapy; OR = odds ratio

## DISCUSSION

In this retrospective cohort of Latino adults with MS, NMOSD, and MOGAD receiving care at a tertiary neuroimmunology clinic, we found that some geographic and socioeconomic factors were associated with clinical outcomes, but not when adjusting for covariates. Nearly half of patients lived ≥50 miles from the UNC MS/Neuroimmunology clinic. In unadjusted analyses, patients living ≥50 miles away had higher EDSS scores than those living closer to the clinic. However, distance to clinic was not independently associated with EDSS after adjustment, suggesting that geographic distance may not be the sole contributor to disability and may be more affected by other clinical and socioeconomic factors rather than as an isolated contributor to disability.

The association between greater distance and higher EDSS in unadjusted analyses may reflect barriers to longitudinal specialty care, including transportation limitations, travel costs, time away from work, delayed evaluation of new symptoms, and difficulty maintaining regular follow-up(Lin et al., 2023; McGinley et al., 2024; Vrtikapa et al., 2025). In one study, Latino individuals with neurologic conditions were 40% less likely to see outpatient neurologists than non-Latino individuals(Saadi et al., 2017). This may contribute to delayed diagnosis, missed disease progression, and fewer opportunities for DMT escalation if one is needed following a relapse or disease progression (McGinley et al., 2024). These barriers may be particularly relevant for patients with demyelinating diseases, for whom periodic ongoing neurologic evaluations for disability progression, MRI surveillance, and DMT escalation are crucial to preventing disease activity and disability accumulation(Montalban et al., 2018). However, the attenuation of the association between distance to the clinic and EDSS after adjustment suggests that geographic distance may be only one of many factors contributing to disability, including disease phenotype, insurance status, treatment access, and individual disease characteristics.

Neighborhood deprivation also had some effect on disability. In our study, the mean ADI state rank was 5.56. Higher ADI state rank was significantly associated with higher EDSS in univariable regression analyses and trended toward significance in multivariable analyses. This effect on disability is consistent with a prior study that found that neighborhood socioeconomic disadvantage was associated with worse neurologic outcomes among people with MS(Abbatemarco et al., 2022). The attenuation of the association after adjustment in our analyses may reflect that other healthcare access-related factors play a role in the association between socioeconomic factors and disability burden. In addition, patients living in high-deprivation neighborhoods with more severe disability may face barriers to reaching tertiary specialty care, potentially leading to underrepresentation of those with highest disability burden in our clinic-based cohort.

We also found that insurance status was significantly associated with both EDSS and ARR in multivariable analyses. Medicaid insurance was associated with higher EDSS and higher ARR compared with commercial insurance in adjusted linear regression models. This finding should not be interpreted as a causal effect of Medicaid itself, but rather as a marker of broader healthcare access-related factors and socioeconomic vulnerability. Insurance status may account for barriers not fully measured by distance or ADI, including prior authorization requirements, medication affordability, DMT approval delays, access to infusion services, and care continuity(Mizell, 2024; Simacek et al., 2018). For example, one prior population-based study of MS found that insurance coverage alone does not ensure access to healthcare, including specific prescription drugs or preferred hospital (Minden et al., 2007). Another study found similar findings as in ours, with Medicaid coverage associated with worse clinical outcomes among patients with MS(Sierra Morales et al., 2025). Among Latino patients specifically, insurance-related barriers might also be associated with Spanish language barriers, health literacy, immigration-related concerns, and difficulty navigating complex health systems(Escobedo et al., 2023; Saadi et al., 2020; Wilson et al., 2005). Many Latino patients seen in clinics rely on certified medical interpreters for communication with their medical provider, which may exacerbate challenges in navigating complex healthcare systems(Escobedo et al., 2023; Wilson et al., 2005). Sociopolitical factors and immigration-related concerns may also contribute to hesitation or reluctance to engage with the medical system(Saadi et al., 2020).

Our finding that ADI state rank was not independently associated with EDSS after adjustment for relevant covariates is consistent with the other studies showing that composite socioeconomic measures do not uniformly predict disability outcomes across all MS cohorts. For example, in a recent study of Latino people with MS, area-level Social Deprivation Index was associated with some disability outcomes in univariable models but was no longer associated with disability after person-centered SDOH factors were included in multivariable models(Amezcua et al., 2025). Our study did not measure person-centered SDOH factors, which may be more individualized predictors of disease burden among Latino patients than area-level indices alone. Similarly, another study found that Black race and Latino ethnicity were not significantly associated with EDSS after adjustment for individual and neighborhood-level SDOH indicators(Orlando et al., 2023), further suggesting that disability disparities may be mediated by person-centered social and structural factors rather than race and ethnicity alone. In another recent study, DMT delay was not significantly associated with EDSS progression after adjustment, despite significant associations with relapses(Sierra Morales et al., 2025), highlighting that healthcare access related factors may impact some MS outcomes more strongly than others. These findings suggest that broad neighborhood-level socioeconomic indices, such as ADI, are useful markers of structural context but may need to be combined with person-centered measures of healthcare access, language barriers, health literacy, and others to better explain disability outcomes in Latino patients with CNS demyelinating disease.

This study has several limitations. This was a single-center study in a tertiary academic center in the U.S. South that included a modest sample size, particularly for NMOSD and MOGAD subgroups, which may limit generalizability across all Latino people with CNS demyelinating diseases. Latino ethnicity was based on electronic medical record documentation of self-reported ethnicity and may be incomplete or inadvertently misclassified. Distance to clinic was based on residential location and does not fully capture travel time, transportation access, availability of local neurology care, telehealth use or access, or ability to attend urgent visits to our clinic for relapse or symptomatic evaluation. ADI reflects neighborhood-level socioeconomic context and may not assess individual-level income nor socioeconomic status, education, employment, documentation status, immigration-related barriers, language preference, or health literacy. Although EDSS and MRI findings were corroborated by a neuroimmunologist based on chart review, relapse history and DMT adherence were abstracted from neuroimmunology office visit note documentation rather than collected using prospective questionnaires or surveys answered by the patient, which may introduce limitations in data accuracy. Moreover, the retrospective nature of this study does not allow causal inferences to be made. In addition, the analysis in this study did not look into patients who have dual insurance (i.e. both Medicaid and Medicare). This study also did not look into DMT duration, or DMT efficacy group as covariates, as DMT duration and type can significantly affect neurological outcomes. Time from the first neurological symptoms to the diagnosis of neurological disease, and time from diagnosis to the start of DMT were also not evaluated as covariates in this study, and both of these variables can be significant contributors to disease activity and disability in MS and related diseases(Chalmer et al., 2018; He et al., 2020). Finally, our study did not assess migration effects (at the time of diagnosis and at the time of onset of neurological symptoms, as some patients might have lived at a different distance from MS specialty care centers).

Despite these limitations, our study provides one of the few studies of Latino adults with MS, NMOSD, and MOGAD in the predominantly rural Southern United States. We found that greater distance to specialty care was associated with higher EDSS in unadjusted analyses, while Medicaid insurance was associated with higher EDSS and ARR in adjusted models. ADI state rank showed a non-significant trend toward higher EDSS, and DMT non-adherence was uncommon among current DMT users. Our findings support the need for prospective studies that incorporate person-centered measures, including transportation burden, insurance barriers, language access, treatment navigation, and others, to better understand and improve outcomes among Latino patients with demyelinating diseases.

## Data Availability

A de-identified dataset may be made available from the corresponding author upon reasonable request and with appropriate institutional approvals.

## Acknowledgements

The authors thank the UNC Carolina Data Warehouse for Health for support with cohort identification and data access.

## Author contributions / CRediT statement

LF: Data curation, investigation, writing – original draft, writing – review and editing.

PR: Data curation, investigation, writing – review and editing.

AM: Formal analysis, methodology, writing – review and editing.

IDB: Conceptualization, supervision, writing – review and editing.

DS: Conceptualization, supervision, writing – review and editing.

MMD: Conceptualization, methodology, supervision, data curation, formal analysis, writing – original draft, writing – review and editing.

## Declaration of generative AI use

The authors used AI to assist with refinement of manuscript text. After using this tool, the authors reviewed and edited the content completely and take full responsibility for the content of the published article.

